# Building Population Phenotypic Journeys from Laboratory Tests in Electronic Health Records for Translational Research

**DOI:** 10.1101/2022.10.10.22280880

**Authors:** Xingmin A Zhang, Kyeryoung Lee, Lan Jin, Zongzhi Liu, Lei Ai, Tomi Jun, Mitch K. Higashi, Qi Pan, William Oh, Gustavo Stolovitzky, Eric Schadt, Peter N. Robinson, Xiaoyan Wang

**Author notes:** Correspondence: Xingmin A Zhang and Xiaoyan Wang.

## Abstract

Abundant volumes of clinical laboratory test results available within Electronic health records (EHRs) are essential for differential diagnosis, treatment monitoring, and outcome evaluation. LOINC2HPO is a recently developed deep phenotyping approach to transform laboratory test results into the Human Phenotype Ontology (HPO) terms. Here, we deployed the approach on a large EHR dataset from the Sema4 Data Warehouse to build patient phenotypic journeys at scale. Among 1.07 billion laboratory test results, we successfully transformed 774 million (72.5%) into HPO-coded phenotypes and built a patient phenotypic journey for over 2.2 million patients. First, a global analysis of patient phenotypic journeys revealed a longitudinal increase in patients with genitourinary system abnormality. The analysis also revealed abnormal phenotypes with strong racial patterns. Second, using severe asthma as an example case, we identified abnormal phenotypes in the past three years that were correlated with asthma progression to severe state. Lastly, we demonstrated that converting laboratory test results into HPO terms resulted in limited information loss. Our study demonstrated that the phenotypic journey framework opens the way to characterize phenotypic trajectories in population level and screen biomarkers for translational research.

## Introduction

Electronic health records (EHRs) have been widely adopted across the world in the past decades. EHRs have been used to build individual patient journeys, which visualize a patient’s progression across the care continuum for a specific disease or condition ^1^. The patient journeys contain narrative timelines of many data elements such as office visits, medical procedures, laboratory tests, diagnoses, and treatments ^2^. Comprehensive patient journeys provide the foundation to identify unmet medical needs for healthcare providers to improve care delivery.

They also reveal the opportunities for the pharmaceutical industry to develop new medical interventions, which may ultimately improve the patient outcome ^1^. The granular “ phenotypes”, which we define in this report as those that describe patient symptoms that deviate from the normal states, are critical components of a patient journey and provide the foundation for numerous topics in clinical-genomic research ^3–5^. There have been previous efforts in extracting patient phenotypes from clinical notes ^6,7^, or mapping patient phenotypes from diagnosis codes ^8^. However, it is still challenging to systematically extract phenotypes, especially at a large scale of population, from EHRs. Developing automated approaches can fasten the extraction of phenotypes from EHRs and support the building of patient journeys for translational research.

Laboratory tests are increasingly identified with Laboratory Observation Identifier Names and Codes (LOINC). For laboratory tests in legacy EHR datasets, retrospectively mapping them from institutional-specific codeset into LOINC can significantly improve interoperability ^9^. Despite this, LOINC codes still have their own limitations when utilized for translational research ^10^. In particular, there are multiple LOINC codes for similar laboratory tests. For example, for urine nitrite measurements, some tests use automated equipment, whereas others use a test strip; some report test values in mg/dL, while others report a binary value (as positive or negative). An abnormal finding in any of the above laboratory tests would indicate nitrituria, but it remains challenging to integrate them with automated inferencing^11^. Therefore, in the era of big data and medicine, it remains extremely challenging to interpret and integrate large scale laboratory tests systematically at the population level, in particular for quantitative tests.

Recently, a computational approach, LOINC2HPO, was developed to semantically integrate clinical laboratory tests by transforming LOINC-coded laboratory test results into the Human Phenotype Ontology (HPO) terms^12^. The HPO is a comprehensive vocabulary that systematically describes and organizes medically relevant abnormal phenotypes. With more than 16,000 terms, the HPO has become the standard ontology for genomic diagnosis of rare diseases and increasingly used in translational research with EHRs ^11,13^. LOINC2HPO includes a mapping library that annotated the outcome of ∼3000 commonly used LOINC-coded laboratory tests into their corresponding HPO terms. Additionally, LOINC2HPO includes an open source software to compare the test value with the normal reference range, and then lookup the mapping library to select the correct HPO term for the observed outcome (i.e. lower than normal, higher than normal, normal etc). In a pilot study, LOINC2HPO was used to generate a list of detailed patient phenotypes from laboratory tests, and identified biomarkers for acute asthma and frequent prednisone usage with a 15,681 patient EHR dataset ^12^.

In the current study, we report building a large collection of patient phenotype journeys from clinical laboratory tests after LOINC2HPO transformation in a large EHR dataset from the Sema4 Data Warehouse (SDW, Figure 1). We mapped over 1 billion laboratory test records from the local test codes to LOINC, and then used the LOINC2HPO approach to transform each laboratory record into the HPO-coded phenotypes. Based on the time course of laboratory tests (LOINC codes) and normal or abnormal test results (HPO terms) over time for each patient, we built longitudinal patient phenotypic journeys for over 2.2 million patients. A global analysis of the patients’ longitudinal journeys revealed changes of tested and observed phenotypes over time and differences among races. Combining with diagnosis records in EHR, we identified abnormal phenotypes, i.e. biomarkers, that were correlated with a progression into severe asthma. Even though transforming continuous values to HPO terms caused information loss in some predictive tasks, we have proved the robustness and interpretability of the patient phenotype journeys resulted from LONIC2HPO. The present study demonstrates that the phenotypic journey framework can be readily implemented and deployed across multiple health systems, which allows systematic biomarker screening and population phenotype analysis for translational research.

**Figure 1.**
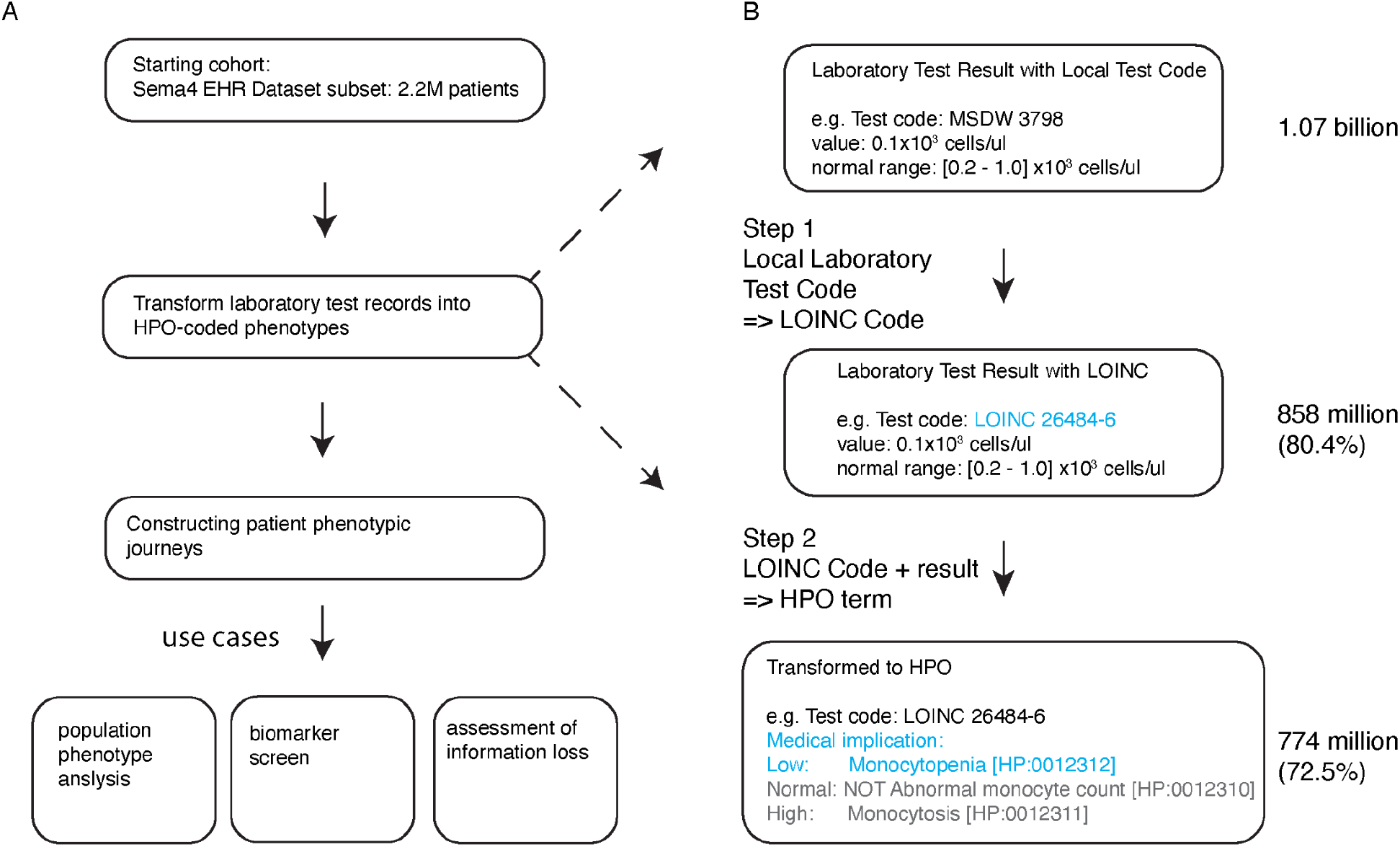
Study design. A. Data analysis steps. We started with 2.2 million patients in Sema4’s EHR dataset. We then transformed laboratory test records for the 2.2 million patients into HPO terms. Using the resulting HPO-coded phenotypes, we constructed a large repertoire of patient phenotypic journeys. We then conducted three translational research analyses to demonstrate the utility of patient phenotypic journeys, including population-level phenotype analysis for longitudinal trends and racial patterns, biomarker screen to identify phenotypes correlated with a future diagnosis of severe asthma and assessment of information loss caused by LOINC2HPO in predictive tasks. B. A two-step process to transform laboratory test results into HPO-coded phenotypes. Step 1, mapping local laboratory test code into LOINC; Step 2, interpret the report laboratory test value and identify the phenotype term for the corresponding LOINC test and actual outcome..

## Methods

### Sema4 Data Warehouse (SDW)

Sema4 is a patient-centered healthcare intelligence company. Sema4 ingested healthcare data from multiple collaborators and maintains a database of EHRs for multi-million patients. The cohort used for this study were from a major health system in the New York area, covering de-identified data from Year 2003 to 2020. We retrospectively enrolled 2.2 million patients whose laboratory test data were recorded longitudinally into this study. This study is exempted from review by the Institutional Review Board (IRB) because the data is de-identified.

### Transforming lab tests in SDW into HPO-coded phenotypes

Laboratory tests were identified with a local codeset in the SDW dataset. We developed a two-step process to transform them into HPO terms (Figure 1B). In the first step, we manually curated a mapping file from the local test codes (n = 11937) into LOINC codes. The mapping process relied on the name, description and unit of the laboratory tests, and matched each to the closest LOINC code. Two curators with biomedical training conducted the process independently, and disagreements were resolved by discussion. With the mapping file, we mapped laboratory test results from the local test code to LOINC. In the second step, we adopted the previously reported LOINC2HPO approach^12^. Briefly, we compared the observed value with the reference range that came with each lab record and interpreted the value as L (lower than normal), H (higher than normal) or N (normal) for Qn (quantitative) laboratory tests, or POS (positive) and NEG (negative) for Ord (Ordinal) laboratory tests. We then utilized the loinc2hpoAnnotation mapping file^12^ to identify the correct HPO term for the corresponding lab code (LOINC) + outcome (L/H/N/POS/NEG).

We additionally conducted ontology-guided inference to add parent terms for each abnormal phenotype transformed from a laboratory test result. For example, for Monocytopenia [HP:0012312] that was transformed from the original laboratory test result (LOINC 26484-6 Monocytes in Blood, 0.1×10^3^ cells/ul, normal range [0.2-1.0]×10^3^ cells/ul), we automatically inferred that the same lab result was also transformed to all the ancestor terms of Monocytopenia [HP:0012312] according to the hierarchy of the HPO, including Abnormal monocyte count [HP:0012310], Abnormal leukocyte count [HP:0011893], Abnormal leukocyte morphology [HP:0001881], Abnormal blood and blood-forming tissues [HP:0001871] and finally Phenotypic abnormality [HP:0000118].

### Building patient journeys from laboratory test-derived phenotypes

We built patient phenotypic journeys for 2.2 million patients using laboratory test-derived phenotypes after the LOINC2HPO transformation and HPO-guided ontology inference. We organized patient phenotype journeys along calendar years (Figure 3). Patient phenotypes include abnormal ones that were derived from laboratory tests with out-of-range values, and normal ones that were derived from those with within-range values. For every abnormal and normal phenotype, we counted how many times it was confirmed by laboratory test results in each calendar year. Because a patient could receive multiple laboratory tests for the same medical problem, it is common to see instances when a patient had a phenotype listed as both “ normal” and “ abnormal”, indicating the laboratory test results yielded different outcomes.

### Longitudinal changes of observed phenotypes and tested phenotypes

At the patient level, a phenotype is considered to be “ observed” if a laboratory test result can directly confirm it. In each calendar year and for each phenotype, we counted the number of patients with at least one laboratory test result that indicated the subject had such an abnormality. Since the database size dramatically increased over time, we normalized the raw patient counts for each observed phenotype by the database size. The database size is defined as the total count of patients who had at least one laboratory test result at each given year.

At the patient level, a phenotype is considered to be “ tested” if a laboratory test can potentially yield a result to confirm whether the subject has such an abnormality, for example, both Hyperglycemia [HP:0003074] and Hypoglycemia [HP:0001943] are tested by fasting glucose measurement (LOINC 1558-6). We systematically collected the information from the loinc2hpoAnnotation library–for each LOINC code, all the mapped HPO terms and their ancestor terms are considered to be “ tested phenotype” by the given LOINC test. For example, LOINC 26484-6 Monocytes in Blood is mapped to three HPO terms, Monocytopenia [HP:0012312] (for lower than normal), Monocytosis [HP:0012311] (for higher than normal) and Abnormal monocyte count [HP:0012310] (negated for normal result). Their ancestors include Abnormal leukocyte count [HP:0011893], Abnormal leukocyte morphology [HP:0001881], Abnormal blood and blood-forming tissues [HP:0001871] and finally Phenotypic abnormality [HP:0000118]. All the above phenotypes are considered to be “ tested” for a subject by the fact of receiving a lab order for LOINC 26484-6 Monocytes in Blood. With this logic, we collected all the laboratory test orders and the tested phenotypes for each order.

Similar to the analysis of observed phenotypes, we counted all the patients who had at least one laboratory test order that indicated the subject had been tested for each HPO-coded phenotype at each given year. Database size is defined the same way as described above, and patient counts are normalized by database size in the corresponding year.

### Racial patterns of observed and tested phenotypes

For each phenotype abnormality, we counted all the patients of each race that were observed (or tested) in each given year. Race was self-reported in our dataset. Additionally, we collected patient counts by race by counting patients that received at least one laboratory test in each year. Then for each phenotype abnormality, we then calculated the odds ratio for each race being observed (or tested) for the corresponding phenotype compared with the other races combined in each year. The procedure generated odds ratios over the years that are indicative of whether a phenotype was being observed or tested compared to other races.

### Screen for biomarkers for progression into severe asthma

The starting cohort was defined as patients who had asthma and asthma-related symptoms by ICD-10 J45 asthma and ICD-10 R06 wheezing. The cohort is further divided into two groups, a severe asthma group by having diagnosis codes of ICD-10 J45.5 severe asthma (n = 3566 patients) and a non-severe asthma group defined as not having diagnosis codes of severe asthma (n = 323,420 patients).

For the severe asthma group, we further identified a subgroup (n = 2593 patients) who had non-severe asthma diagnoses before severe asthma, indicating that the cohort progressed from non-severe asthma to severe asthma. The first severe asthma diagnosis of such patients was defined as t_0_. And all phenotypes derived from laboratory tests with an abnormal finding in the three years immediately prior to t_0_ were collected and used for the biomarker screening.

For the non-severe asthma group, we additionally excluded 1% patients who had mentions of “ severe asthma” in their clinical notes. We randomly sampled a subset (2x of patients who progressed to severe asthma) as the controls for our data analysis. The latest asthma diagnosis was defined as t_0_ for these patients. Similarly, all the laboratory test-derived phenotypes in the previous three years were collected for the biomarker screening.

### Evaluation of information loss caused by converting continuous laboratory test data into HPO terms

We compared the performance of using original laboratory test values vs the transformed HPO-coded phenotype to predict future disease diagnoses. Table 1 listed the selected diseases, case/control definitions and the corresponding laboratory tests. Briefly, we selected four disease diagnoses; abnormal liver function, acute kidney failure, colorectal cancer and aplastic anemia. For abnormal liver function, we selected a cohort of patients by diagnosis codes, ICD10 R94.5 or ICD9 794.8: Abnormal results of liver function studies, as cases. If a patient had multiple records of the diagnosis codes, the earliest one was chosen for the patient. For each patient and each LOINC test, we used the most recent record prior to the diagnosis as a feature. The same set of patients served as their own controls as we reasoned the diagnosis of abnormal liver function was transient and thus it was reasonable to assume the patients had a healthy liver function 180 days ago. We chose the 180 day cutoff because we observed laboratory test values beginning to trend up/down after this date (Supplemental Figure 3). For the controls, we selected the latest laboratory test at least 180 days prior to the diagnosis as the feature. After setting up cases/controls and collecting laboratory test values, we built a logistic regression classifier to predict whether the patient is case/control from each laboratory test, either using the original numeric value (e.g. gamma glutamyl transferase, 91 u/L) or the transformed HPO phenotype (binary, e.g. Elevated gamma-glutamyltransferase level [HP:0030948]: observed or not). A random split (70%) of data was used for training the classifier and the remaining 30% was used as the testing data. We built the receiver operating characteristic (ROC) curve from the predictions/labels of testing data and calculated the area under curve (AUC) as the performance metric.

**Table 1.** disease diagnoses and LOINC tests for comparing numeric vs HPO-coded phenotypes in predictive tasks

| diagnosis | cohort definition | LOINC test |
| --- | --- | --- |
| abnormal liver function | case: ICD-10 R94.5 or ICD-9 794.8;<br>control: same patients at 180 days prior to the diagnosis of abnormal liver function | LOINC 1742-6 Alanine aminotransferase [Enzymatic activity/volume] in Serum or Plasma |
|  |  | LOINC 1920-8 Aspartate aminotransferase [Enzymatic activity/volume] in Serum or Plasma |
|  |  | LOINC 6768-6 Alkaline phosphatase [Enzymatic activity/volume] in Serum or Plasma |
|  |  | LOINC 2324-2 Gamma glutamyl transferase [Enzymatic activity/volume] in Serum or Plasma |
|  |  | LOINC 1968-7 Bilirubin.direct [Mass/volume] in Serum or Plasma |
|  |  | LOINC 1975-2 Bilirubin.total [Mass/volume] in Serum or Plasma |
|  |  | LOINC 1751-7 Albumin [Mass/volume] in Serum or Plasma |
|  |  | LOINC 2885-2 Protein [Mass/volume] in Serum or Plasma |
|  |  | LOINC 2532-0 Lactate dehydrogenase [Enzymatic activity/volume] in Serum or Plasma [DISCOURAGED] |
|  |  | LOINC 14805-6 Lactate dehydrogenase [Enzymatic activity/volume] in Serum or Plasma by Pyruvate to lactate reaction |
|  |  | LOINC 5902-2 Prothrombin time (PT) |
| acute kidney failure | case: ICD-10 N17 as the primary diagnosis;<br>control: same patients at 100 days prior to the diagnosis of acute kidney failure (100 days ago) | LOINC: 2160-0 Creatinine [Mass/volume] in Serum or Plasma |
|  |  | LOINC: 3094-0 Urea nitrogen [Mass/volume] in Serum or Plasma |
|  |  | LOINC: 6299-2 Urea nitrogen [Mass/volume] in Blood |
|  |  | LOINC: 2823-3 Potassium [Moles/volume] in Serum or Plasma |
|  |  | LOINC: 5804-0 Protein [Mass/volume] in Urine by Test strip |
|  |  | LOINC: 2889-4 Protein [Mass/time] in 24 hour Urine |
|  |  | LOINC: 2888-6 Protein [Mass/volume] in Urine |
| aplastic anemia | case: ICD-10 D61.9 or ICD-9 284.9 ;<br>control: same patients when they were healthy (100 days ago) | LOINC: 789-8 Erythrocytes [# /volume] in Blood by Automated count |
|  |  | LOINC: 6690-2 Leukocytes [# /volume] in Blood by Automated count |
|  |  | LOINC: 777-3 Platelets [# /volume] in Blood by Automated count |
|  |  | LOINC: 751-8 Neutrophils [# /volume] in Blood by Automated count |
| colorectal cancer | case: ICD-10 C18.0-9, C19.9 or C20.9, excluding patients with any other cancer diagnosis prior to colorectal cancer;<br>control: sex, race and age-matched patients free from any cancer, including ICD-9 140, 172, 174, 209, 225, 227, 228, 230, 234, 237, 238, 239, 273, 277 and ICD-10 C00, C43, C4A, C45, C48, C49, D00, D09, D18, D32, D33, D35, D42, D43, D44, D45, D46, D49, D85, D87 | LOINC: 6690-2 Leukocytes [# /volume] in Blood by Automated count |
|  |  | LOINC: 789-8 Erythrocytes [# /volume] in Blood by Automated count |
|  |  | LOINC: 718-7 Hemoglobin [Mass/volume] in Blood |
|  |  | LOINC: 4544-3 Hematocrit [Volume Fraction] of Blood by Automated count |
|  |  | LOINC: 787-2 MCV [Entitic volume] by Automated count |
|  |  | LOINC: 785-6 MCH [Entitic mass] by Automated count |
|  |  | LOINC: 786-4 MCHC [Mass/volume] by Automated count |
|  |  | LOINC: 21000-5 Erythrocyte distribution width [Entitic volume] by Automated count |
|  |  | LOINC: 788-0 Erythrocyte distribution width [Ratio] by Automated count |
|  |  | LOINC: 777-3 Platelets [# /volume] in Blood by Automated count |
|  |  | LOINC: 32207-3 Platelet distribution width [Entitic volume] in Blood by Automated count |
|  |  | LOINC: 32623-1 Platelet mean volume [Entitic volume] in Blood by Automated count |

We adopted a very similar approach to analyze laboratory tests for acute kidney failure and aplastic anemia. For acute kidney failure, cases were defined as patients who had a primary diagnosis of ICD10 N17 Acute kidney failure. For aplastic anemia, cases were defined as patients who had a primary or secondary diagnosis of ICD-10 D61.9 or ICD-9 284.9 Aplastic anemia, unspecified. For both diseases, we chose 100 day as the cutoff to designate patients as healthy controls of themselves, as we observed laboratory tests beginning to change at around this time (Supplemental Figure 4 and 5).

For colorectal cancer, cases were defined as patients who had a primary diagnosis of ICD-10 C18, C19, C20, or ICD-9 153, 154.0, 154.1, or 154.8. For patients with multiple diagnoses, the earliest one was chosen. Controls were defined as patients who were free from any cancer diagnoses (based on diagnosis codes ^14^; see Table 1 for details). We sampled controls by matching them 1:1 to cases based on sex, age and race (Supplemental Figure 6). For each patient and each LOINC test, we selected the most recent record prior to colorectal cancer diagnosis (for cases) or the latest record in the history (for controls) as the features (Supplemental Figure 7). Statistical analysis was performed in the same manner as described in liver function analysis.

### Code and statistical analyses

Raw EHR data was hosted on an Amazon Redshift cluster. Data transformations, including preprocessing of EHR data and the LOINC2HPO transformation, were conducted by SQL within the database cluster and managed by a custom Java application. Summary statistics collected from the Redshift cluster were loaded into memory and further analyzed in R. The interactive web application that allows user to explore the longitudinal and racial differences of more granular HPO phenotypes, Sema4 Lab Phenotype Viewer, was coded with RShiny. All statistical analysis was conducted in R (version 4.1.2).

## Results

### SDW cohort description

The SDW EHR dataset contains medical records for over 12 million patients. We focused on a subset of patients from a New York area health system who had at least one laboratory test result in their medical history. The subset has around 2.2 million patients in total. Of these, 56.0% are female and 43.9% are male (Figure 2A). Most patients reported white racial identity (38.5%), followed by Black or African American (16.7%) and Asian (2.7%) (Figure 2B). A small fraction (6.4%) of patients reported different races at different visits and are considered to have a mixed race. In addition, race was not reported for a substantial fraction (29.0%). The cohort has almost equal distributions in the 30-40, 40-50, 50-60, 60-70 and >70 year age brackets, but much less for the under 30s (Figure 2C).

**Figure 2.**
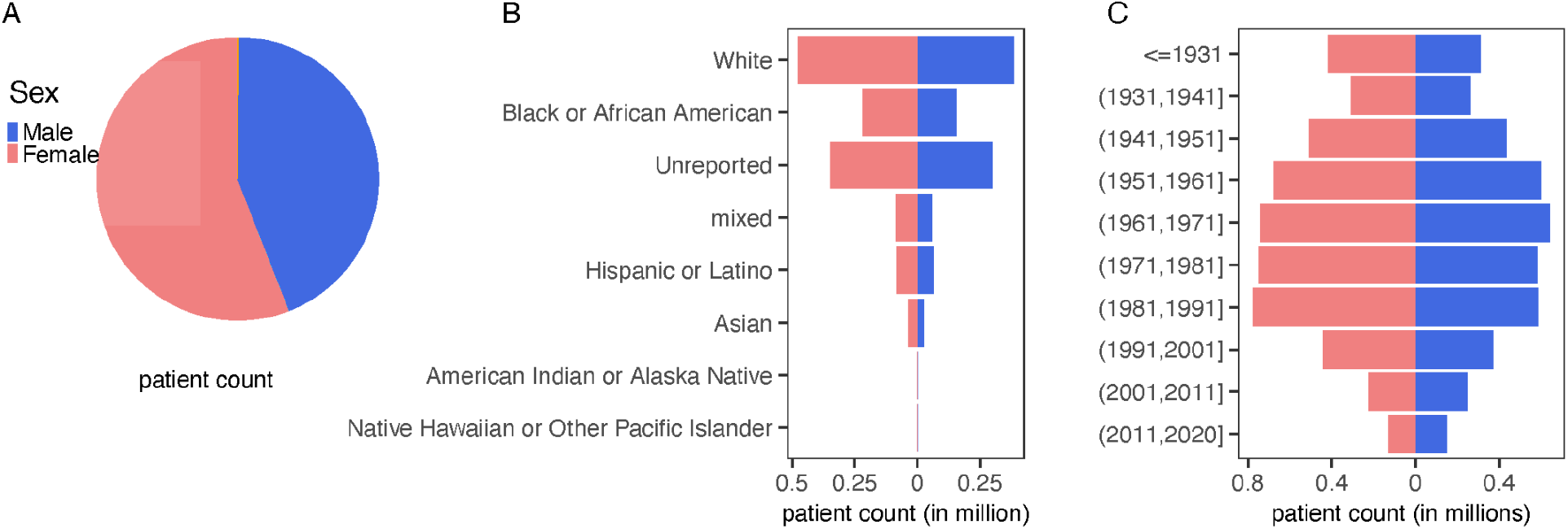
Study demographics. A. Patient distribution based on sex. B. Patient distribution based on self-reported race. C. Patient distribution based on birth year.

### Implementation of LOINC2HPO with SDW data and summary statistics

Because laboratory tests in our dataset were identified with an institution-specific codeset, we first mapped the local codeset to LOINC and then utilized the LOINC2HPO approach ^12^ to transform the lab findings into HPO-coded phenotype terms (Figure 1B). Among 1.07 billion laboratory test records in SDW, we successfully mapped 858 million (80.4%) from local test code into LOINC (Step 1, Figure 1B), and subsequently transformed 774 million (72.5%) to HPO terms. On average, each patient has 324 lab results that are transformed into HPO-coded phenotypes, many of which could be the same kind of laboratory tests that were ordered multiple times during patient care. After removing duplicates, each patient on average has 33.1 phenotypes (Figure 3), including 8.5 abnormal phenotypes from laboratory tests with out-of-range values and the rest are normal phenotypes from laboratory tests with in-range values. Using the hierarchy of HPO, we further inferred 22.1 abnormal phenotypes, i.e. the ancestors of more granular phenotypes directly confirmed by laboratory test results, based on those 8.5 abnormal phenotypes.

**Figure 3.**
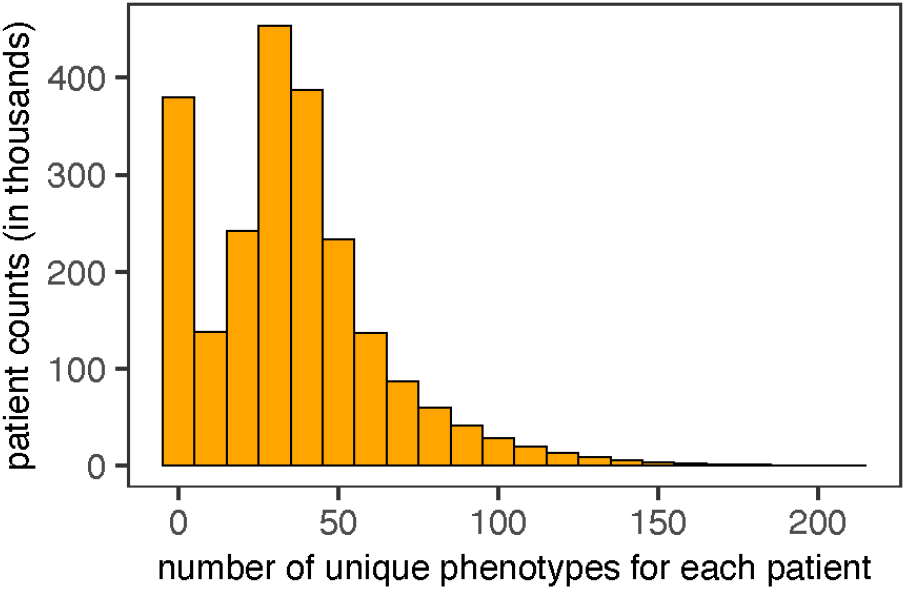
Patient distribution by the number of unique phenotypes throughout their medical history.

### Example patient phenotype journey

With the laboratory test-derived phenotypes and the longitudinal information of their underlying source record, we were able to build patient phenotypic journeys for each of the 2.2 million patients in the cohort. Figure 4 showed a representative example for a middle-aged, female African American patient. The patient journey includes abnormal phenotypes that were directly confirmed by a laboratory test result or inferred based on HPO hierarchy, and normal phenotypes from normal laboratory test values. The example patient was persistently observed to have Elevated serum alanine aminotransferase [HP:0031964] and Elevated serum aspartate aminotransferase [HP:0031956]; the patient, however, did not have Abnormality of alkaline phosphatase level [HP:0004379], Abnormal albumin level[HP:0012116], nor Increased total bilirubin [HP:0003573]. Collectively, the phenotypes indicate that the patient possibly had chronic hepatocellular injury ^15^, which was validated by the repeated assignments of diagnosis codes for fatty liver between 2015 and 2020. In summary, HPO phenotypes transformed from laboratory test results allow one to build patient phenotypic journeys at a large scale and can facilitate differential diagnoses.

**Figure 4.**
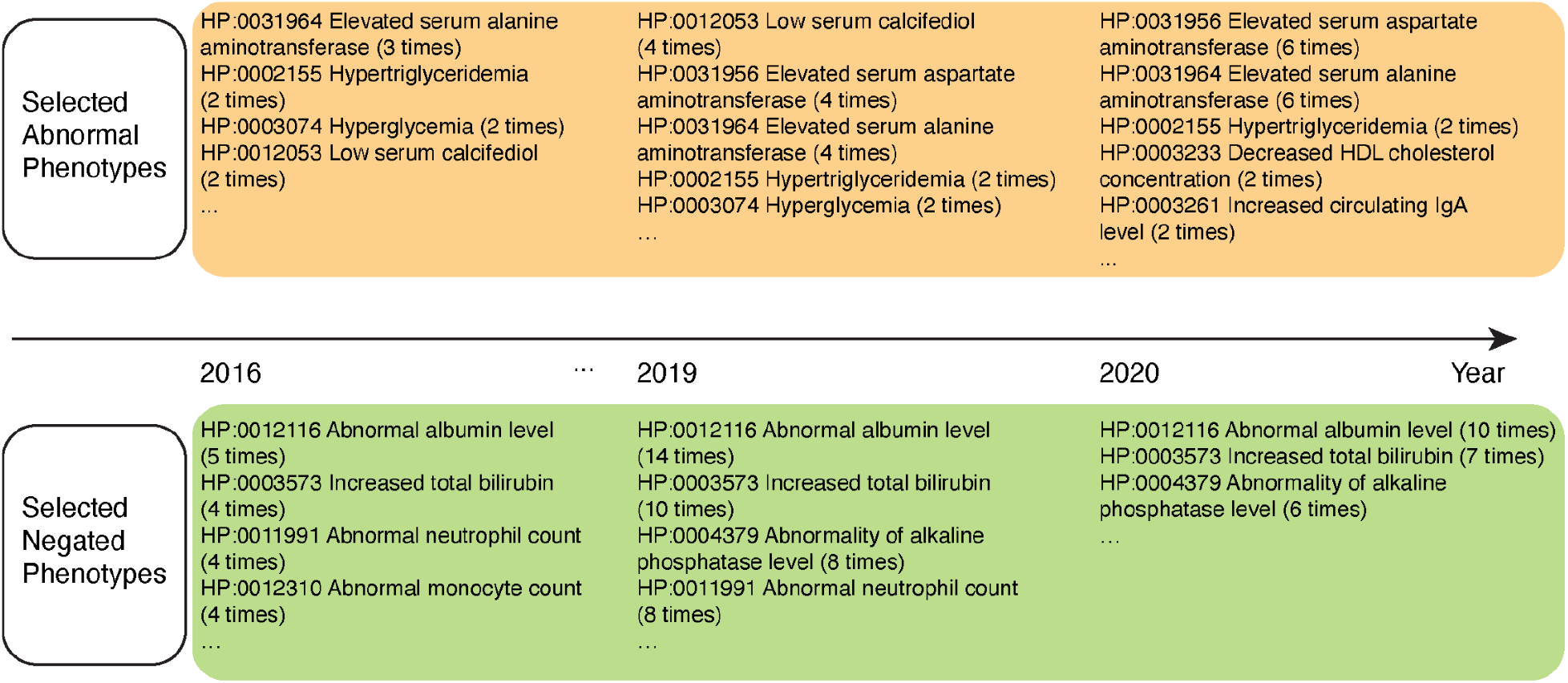
Example patient journey for an African American female subject born in 1967.

### Longitudinal changes of patient counts for observed or tested phenotypes

We conducted a global analysis of 2.2 million patient phenotypic journeys to identify what phenotypes are more or less frequently observed over time. We focused on abnormal phenotypes that were transformed from laboratory tests with out-of-range values and defined a patient to have been observed for an abnormal phenotype as long as there was one laboratory test result to indicate so. Patient counts for all of the high-level HPO terms increased over time (Figure 5A), which was expected as the dataset had a growing number of patients. After normalizing patient counts by the database size in the corresponding year (Figure 5B, Table 2A), we found there were statistically significant decreases in patients observed for Abnormality of blood and blood-forming tissues [HP:0001871], Abnormality of the immune system [HP:0002715], Abnormality of the digestive system [HP:0025031] and Abnormality of the cardiovascular system [HP:0001626]. The decrease for Abnormality of blood and blood-forming tissues [HP:0001871] and Abnormality of the immune system [HP:0002715] was preserved even after taking into consideration how many patients were actually tested (Supplemental Figure 1A), which indicates the cohort was healthier in these categories possibly due to more healthier patients undergoing routine screenings. On the contrary, there are statistically significant increases in patients observed for the Abnormality of the genitourinary system [HP:0000119] and Abnormality of metabolism/homeostasis [HP:0001939]. While the increase for the Abnormality of metabolism/homeostasis [HP:0001939] was explained by more patients undergoing testing, the increase for the Abnormality of the genitourinary system [HP:0000119] was not, indicating that the cohort was less healthier for the genitourinary system (Supplemental Figure 1B). We additionally developed an interactive web application, Sema4 Lab Phenotype Viewer, that allows users to explore the longitudinal changes of more granular phenotype terms (url: https://permanent.link/to/loinc2hpo/sema4_msdw). Overall, the global analysis of observed phenotypes revealed significant cohort changes over time.

**Figure 5.**
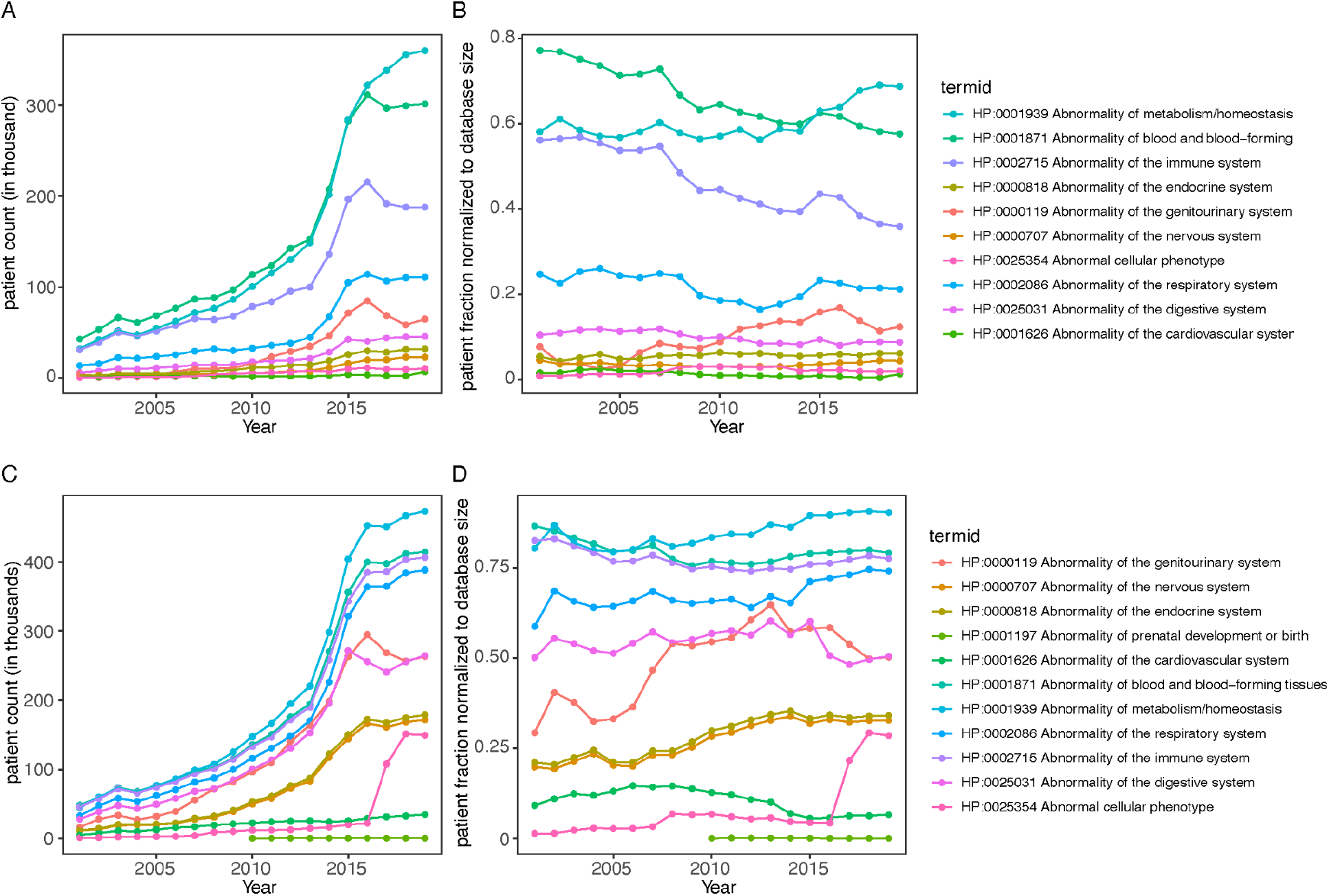
Longitudinal changes of observed (Panel A and Panel B) and tested (Panel C and Panel D) phenotypic abnormalities. A. Raw patient count for those who were observed to have indicated abnormal phenotype. B. Normalized patient count by database size for those who were observed to have indicated abnormal phenotypes. C. Raw patient count for those who were tested for the indicated abnormal phenotype. D. Normalized patient count by database size for those who were tested for the indicated phenotype.

**Table 2A.** Statistical testing for trendline changes of observed phenotypes

| termid | label | beta | p.value | p.value.bonferroni | significance |
| --- | --- | --- | --- | --- | --- |
| HP:0001871 | Abnormality of blood and blood-forming tissues | -1.16E-02 | 2.22E-11 | 2.22E-10 | *** |
| HP:0002715 | Abnormality of the immune system | -1.28E-02 | 6.90E-11 | 6.90E-10 | *** |
| HP:0025031 | Abnormality of the digestive system | -1.82E-03 | 9.79E-06 | 9.79E-05 | *** |
| HP:0000119 | Abnormality of the genitourinary system | 5.85E-03 | 1.91E-05 | 1.91E-04 | *** |
| HP:0001626 | Abnormality of the cardiovascular system | -6.93E-04 | 1.05E-03 | 1.05E-02 | * |
| HP:0001939 | Abnormality of metabolism/homeostasis | 4.41E-03 | 2.48E-03 | 2.48E-02 | * |
| HP:0000818 | Abnormality of the endocrine system | 4.74E-04 | 1.22E-02 | 1.22E-01 |  |
| HP:0002086 | Abnormality of the respiratory system | -2.41E-03 | 2.16E-02 | 2.16E-01 |  |
| HP:0025354 | Abnormal cellular phenotype | 5.59E-04 | 5.58E-02 | 5.58E-01 |  |
| HP:0000707 | Abnormality of the nervous system | 1.19E-04 | 5.51E-01 | 5.51E+00 |  |

Because the orders of laboratory tests typically reflect the medical context, we collected what phenotypes can be theoretically revealed by each laboratory test order, aka. tested phenotype, and determined whether they were changed over time. For high level abnormal phenotypes, there are statistically significant increases in patients tested for Abnormality of the nervous system [HP:0000707], Abnormality of the endocrine system [HP:0000818], Abnormality of cellular phenotype [HP:0025354], Abnormality of the respiratory system [HP:0002086] and Abnormality of the genitourinary system [HP:0000119]; there are statistically significant decreases in patients tested for Abnormality of the cardiovascular system [HP:0001626], Abnormality of the immune system [HP:0002715], and Abnormality of blood and blood-forming tissues [HP:0001871] (Figure 5CD, Table 2B). Similarly, many granular phenotypes are also tested differently and can be explored with the interactive web application, Sema4 Lab Phenotype Viewer. These findings indicate that our patient phenotypic journeys were able to reveal epidemiological trends on what phenotypic tests were ordered more or less frequently over time.

**Table 2B.** Statistical testing for trendline changes of tested phenotypes

| termid | label | beta | p.value | p.value.bonferroni | significance |
| --- | --- | --- | --- | --- | --- |
| HP:0000707 | Abnormality of the nervous system | 7.49E-03 | 5.49E-06 | 6.04E-05 | *** |
| HP:0000818 | Abnormality of the endocrine system | 7.70E-03 | 5.90E-06 | 6.49E-05 | *** |
| HP:0025354 | Abnormal cellular phenotype | 1.14E-02 | 1.12E-04 | 1.23E-03 | ** |
| HP:0001626 | Abnormality of the cardiovascular system | -4.10E-03 | 2.51E-04 | 2.77E-03 | ** |
| HP:0002715 | Abnormality of the immune system | -3.73E-03 | 1.75E-03 | 1.93E-02 | * |
| HP:0001871 | Abnormality of blood and blood-forming tissues | -4.07E-03 | 1.90E-03 | 2.09E-02 | * |
| HP:0002086 | Abnormality of the respiratory system | 4.32E-03 | 2.20E-03 | 2.42E-02 | * |
| HP:0000119 | Abnormality of the genitourinary system | 1.09E-02 | 4.37E-03 | 4.81E-02 | * |
| HP:0001939 | Abnormality of metabolism/homeostasis | 4.16E-03 | 4.75E-03 | 5.22E-02 | . |
| HP:0001197 | Abnormality of prenatal development or birth | -8.10E-05 | 3.66E-02 | 4.03E-01 |  |

### Racial patterns in observed and tested phenotypes

Race is an important factor in clinical observations and translational research using EHR ^16–18^. We analyzed each phenotype to determine whether it was tested and observed, respectively, more frequently in one race compared to other races. Many phenotypes are tested differently in each race. At the high level, Asian were more likely to be tested for Abnormality of the cardiovascular system [HP:0001626] but less likely for Abnormality of prenatal development or birth [HP:0001197] or Abnormal cellular phenotype [HP:0025354] (Figure 6B). Black or African American were more likely to be tested for Abnormality of prenatal development or birth [HP:0001197], but less likely for Abnormal cellular phenotype [HP:0025354], Abnormality of the endocrine system [HP:0000818] and Abnormality of the nervous system [HP:0000707], especially after 2010. White were more likely to be tested for Abnormal cellular phenotype [HP:0025354], Abnormality of the endocrine system [HP:0000818], Abnormality of the nervous system [HP:0000707], and Abnormality of the digestive system [HP:0025031], but less likely for Abnormality of prenatal development or birth [HP:0001197] and Abnormality of the cardiovascular system [HP:0001626].

**Figure 6.**
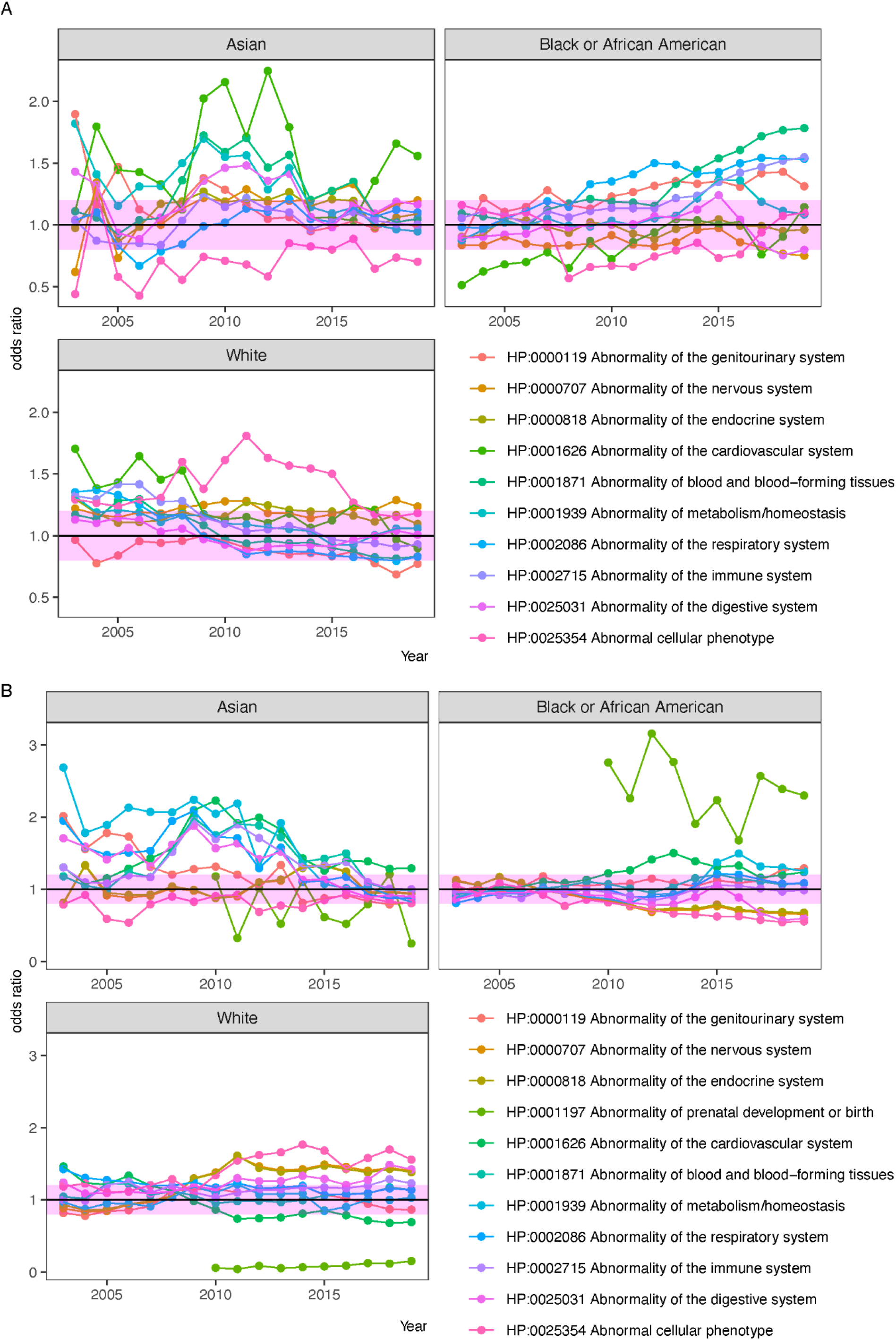
Racial difference of observed phenotypes (A) and tested phenotypes (B). Only high level HPO terms are shown. For other phenotypes, refer to the interactive app at https://permanent.link/to/loinc2hpo/sema4_msdw.

When all the phenotypes were ranked together, we found the most overly tested phenotype in Black or African American were for CD4 ^+^ T helper cells (Table 3), which is consistent with heavier HIV burden in the black population ^19^. Similarly, Positive urine methadone test [HP:0031841] and Positive urine barbiturate test [HP:0500109] are also more likely to be tested in the black population, indicating that the Black population is more likely to be tested for drug abuse. Additionally, the Black population is also more likely to be tested for Persistence of hemoglobin F, which is possibly for the diagnosis of sickle cell anemia, a disease with higher incidence in the black population ^20,21^. The White population were most disproportionately frequently tested for phenotypes of granulocytes, including Granulocytopenia (HP:0001913) and Granulocytosis (HP:0032310), a group of immune cells that are responsible for many autoimmune diseases ^22^. The White population were also much more likely to be tested for very low density lipoprotein (VLDL) cholesterol concentration, which is possibly due to a combined effect of medical needs (e.g. higher incidence of familial hypercholesterolemia) and higher awareness and/or willingness for preventative screenings. In the Asian population, the most likely tested phenotype is Phenotypic abnormality [HP:0000118], which is the most generic phenotype term in HPO. Because Phenotypic abnormality [HP:0000118] can be tested by any laboratory test, the finding suggests that Asian were more likely to receive laboratory tests in general compared with other races in the dataset.

**Table 3.** Top over-proportionally tested phenotypes for each race. OR: odds of having received lab tests for the phenotype in the race vs other races; phenotypes with less than 1700 tested patients were excluded.

| Black or African American |  |  |  |  |
| --- | --- | --- | --- | --- |
| *termid* | *label* | *OR* | *Lab.Record* | *Patient.Count* |
| HP:0002843 | Abnormal T cell morphology | 2.56 | 334106 | 44404 |
| HP:0005403 | Decrease in T cell count | 2.56 | 334106 | 44404 |
| HP:0005407 | Decreased proportion of CD4-positive helper T cells | 2.56 | 311716 | 44404 |
| HP:0011839 | Abnormal T cell count | 2.56 | 334106 | 44404 |
| HP:0025540 | Abnormal T cell subset distribution | 2.56 | 334106 | 44404 |
| HP:0500267 | Abnormal proportion of CD4-positive helper T cells | 2.56 | 311716 | 44404 |
| HP:0031841 | Positive urine methadone test | 2.19 | 35696 | 21433 |
| HP:0011904 | Persistence of hemoglobin F | 2.18 | 27080 | 13461 |
| HP:0500109 | Positive urine barbiturate test | 2.16 | 48469 | 27316 |
| HP:0031961 | Abnormal serum anion gap | 2.16 | 682639 | 206481 |
| White |  |  |  |  |
| HP:0001913 | Granulocytopenia | 16.11 | 129756 | 92956 |
| HP:0032310 | Granulocytosis | 16.11 | 129756 | 92956 |
| HP:0003362 | Increased VLDL cholesterol concentration | 7.14 | 244745 | 197169 |
| HP:0031243 | Decreased VLDL cholesterol concentration | 7.14 | 244745 | 197169 |
| HP:0031889 | Abnormal VLDL cholesterol concentration | 7.14 | 244745 | 197169 |
| HP:0001900 | Increased hemoglobin | 2.17 | 263958 | 12745 |
| HP:0001901 | Polycythemia | 2.17 | 263958 | 12745 |
| HP:0008348 | Decreased circulating IgG2 level | 1.78 | 5992 | 3288 |
| HP:0032135 | Decreased circulating IgG subclass level | 1.78 | 23597 | 3492 |
| HP:0032137 | Decreased circulating IgG3 level | 1.78 | 5975 | 3278 |
| Asian |  |  |  |  |
| HP:0000118 | Phenotypic abnormality | 2.14 | 16291266 | 95861 |
| HP:0032232 | Increased circulating creatine kinase MB isoform | 2 | 17174 | 4036 |
| HP:0001939 | Abnormality of metabolism/homeostasis | 1.89 | 6485715 | 88950 |
| HP:0006254 | Elevated alpha-fetoprotein | 1.69 | 11213 | 3269 |
| HP:0012337 | Abnormal homeostasis | 1.69 | 1358581 | 82468 |
| HP:0003113 | Hypochloremia | 1.64 | 404993 | 69973 |
| HP:0011422 | Abnormal blood chloride concentration | 1.64 | 404993 | 69973 |
| HP:0011423 | Hyperchloremia | 1.64 | 404993 | 69973 |
| HP:0010927 | Abnormal blood inorganic cation concentration | 1.58 | 510106 | 68980 |
| HP:0010930 | Abnormal blood monovalent inorganic cation concentration | 1.57 | 745644 | 71488 |

**Table 4.**
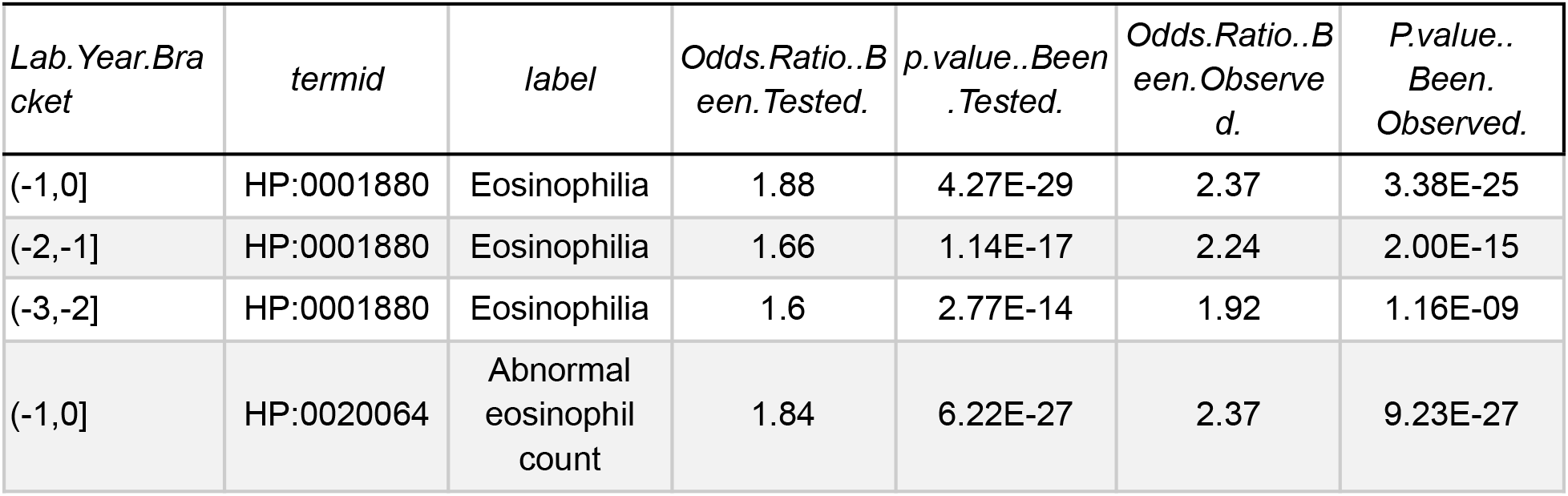
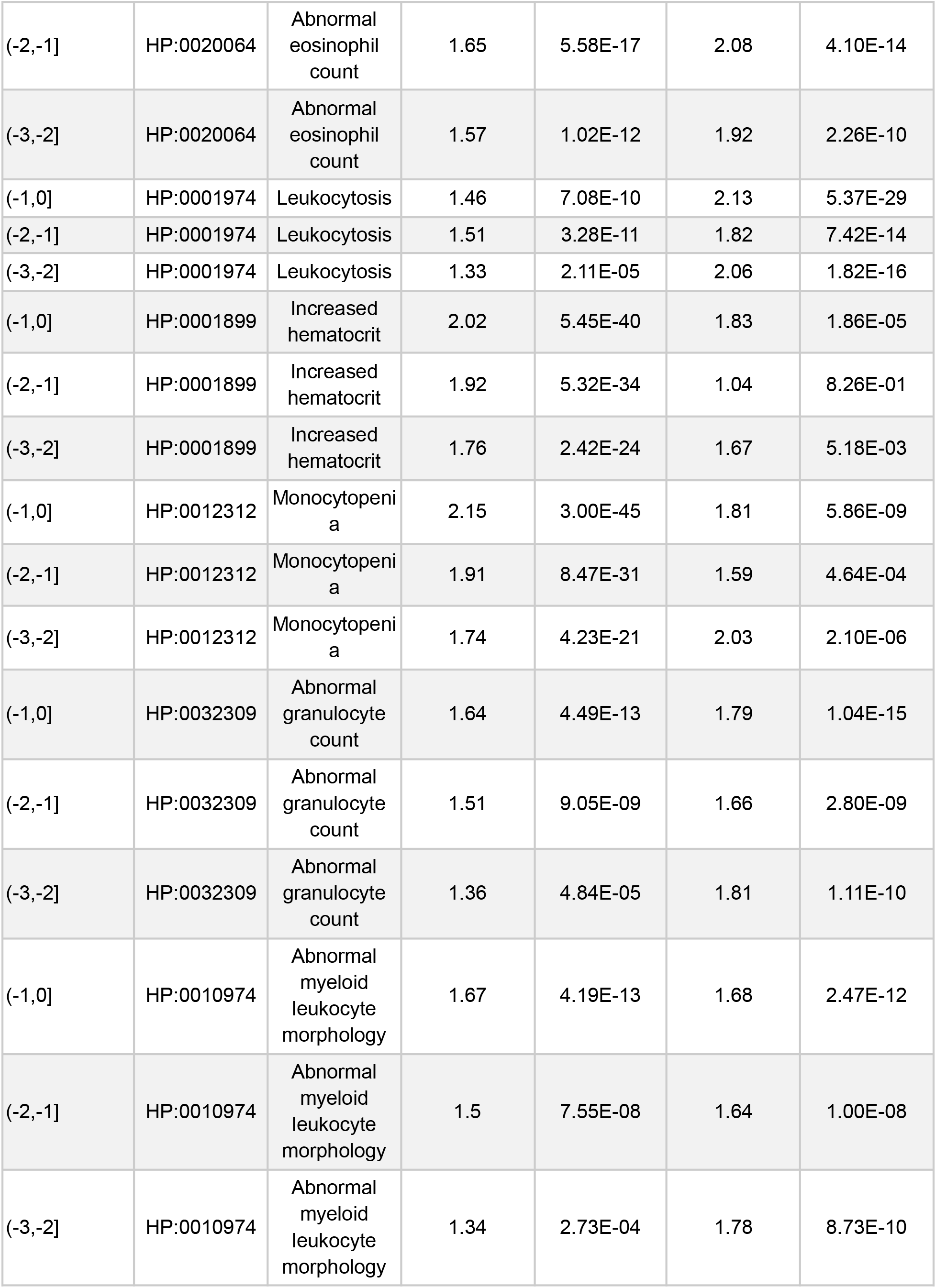
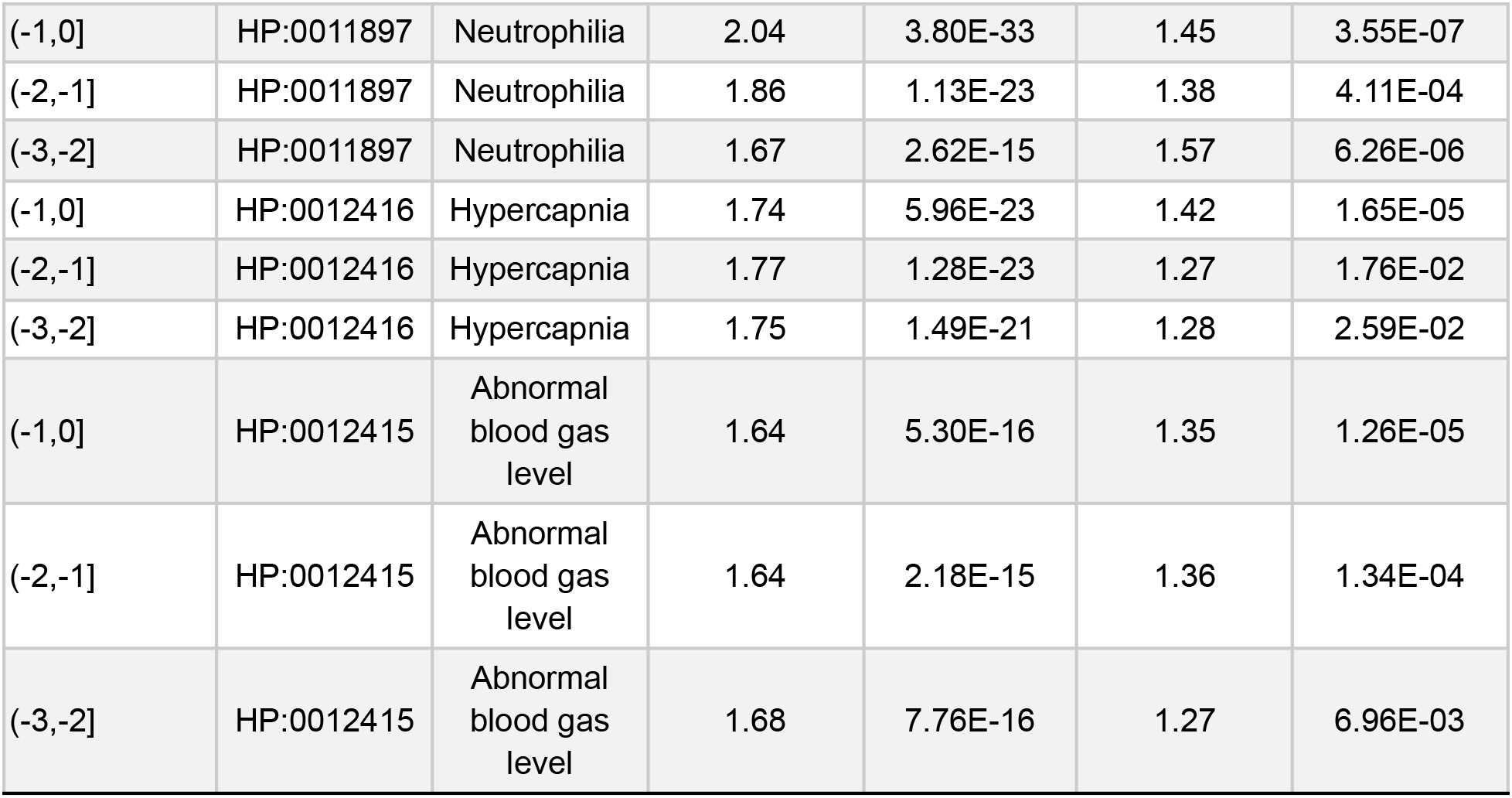
Selection of biomarkers that are statistically significantly correlated with future progression into severe asthma.

When we looked at racial distribution of patients that were observed for each phenotype, we also found apparent differences for many phenotypes (Figure 6A). For example, Asian were consistently more likely to have confirmed Abnormality of the cardiovascular system [HP:0001626], but much less likely for Abnormal cellular phenotype [HP:0025354]. Black or African American were more likely to have Abnormality of the respiratory system [HP:0002086], Abnormality of the genitourinary system [HP:0000119] and Abnormality of blood and blood-forming tissues. White are more likely to have confirmed Abnormal cellular phenotype [HP:0025354] but overall proportionally observed for most phenotypes. However, the differences diminished when patient counts were normalized to the number of patients actually tested for each phenotype(Supplemental Figure 2), suggesting that the racial differences were mainly due to different races being tested differently rather than one race being sicker than others. Taken together, the above findings revealed racial patterns in regards to what medical phenotypes were tested in medical care.

### Screen for biomarkers correlated with progression into severe asthma

To demonstrate how the patient journeys can be used for translational research, we conducted a case study to identify biomarkers associated with asthma progression to the severe form. Asthma is a common condition that affected 25 million (7.8%) of the US population in 2019 ^23^. An estimated 10% of asthma patients develop severe asthma ^24^, which has a significantly higher frequency of exacerbation and can lead to death if not treated in a timely fashion ^25^. Therefore, identifying biomarkers from routinely observed medical phenotypes that can predict whether a patient will progress into severe asthma has significant medical implications. We identified 2593 severe asthma patients who progressed from the non-severe form to the severe form, and twice the number of patients who continued to have non-severe asthma as controls. For each phenotype in the preceding years, we assessed whether testing and observation of the phenotype is correlated with asthma progression into the severe form after adjusting for race, sex and age. We included whether a phenotype is tested or not as an independent variable as we reasoned that true biomarkers should be more likely to be ordered when severe asthma was suspected.

We found that among phenotypes in the year preceding to progression to severe asthma, having been tested for Eosinophilia [HP:0001880] was associated with significantly increased odds for severe asthma progression; in addition, confirmation of Eosinophilia was also strongly correlated with increased odds for severe asthma diagnosis. In a similar manner, Neutrophilia [HP:0011897], which is elevated counts of neutrophils, was also identified to be correlated with severe asthma. Elevated counts of eosinophils and neutrophils in the sputum were identified as important subtypes of asthma ^26^; previous studies also indicate that these cell types in the peripheral blood, are weaker but significant biomarkers for severe asthma ^27–29^. Our findings are consistent with the previous reports, thus validating our approach here. We also identified Leukocytosis [HP:0001974], Abnormal granulocyte count [HP:0032309] and Abnormal myeloid leukocyte morphology [HP:0010974] that are significantly correlated with severe asthma, which are expected as they are the parent terms of Eosinophilia [HP:0001880] and Neutrophilia [HP:0011897]. Interestingly, we identified Monocytopenia [HP:0012312], i.e. low counts of monocytes, as strongly associated with progression to severe asthma. Additionally, we found Increased hematocrit [HP:0001899], Hypercapnia [HP:0012416] and Abnormal blood gas level [HP:0012415] (the parent of Hypercapnia) were strongly associated with severe asthma. Hypercapnia [HP:0012416], i.e. an elevated level of CO_2_ in the blood could be related to the effects of asthma on respiration ^30^; and Increased hematocrit [HP:0001899] could be related to chronic hypoxia, although our analysis does not allow any conclusions about the pathomechanisms of observed laboratory abnormalities..

To assess whether phenotypic abnormalities even further ahead of progression to severe asthma were correlated with the outcome, we conducted similar analysis for phenotypes collected in the one year or two years window before severe asthma diagnosis. We found the same phenotypes also have significant correlations with the later progression into severe asthma, thus suggesting the observed correlations are persistent. In addition, the weights of correlations for most phenotypes gradually decreased as we moved the time window further away from the outcome.

### Assessment of information loss caused by binning numeric laboratory test values

Converting continuous values of laboratory tests to HPO-coded phenotypes are expected to cause information loss, but it is unclear to what extent the loss is. We determined this by comparing the performance of laboratory tests in predicting medical diagnoses when used as the original continuous values vs HPO-coded phenotypes. We selected four representative diseases and laboratory tests that are used for their diagnoses (Table 1). The medical conditions, including abnormal liver function, acute kidney failure, aplastic anemia and colorectal cancer, were chosen so that the laboratory tests cover blood work and urinalysis, which are commonly ordered and utilized for healthcare machine learning tasks ^31,32^. We built logistic regression classifiers to predict whether a patient was a case or control from laboratory tests conducted before the disease diagnosis, one laboratory test at a time. We found that using the transformed HPO-coded phenotype (as a binary value to indicate whether an abnormal phenotype was observed or not) almost always reduced the AUC of ROC (Figure 7), validating the expectation that transforming continuous laboratory tests into HPO caused information loss in predictive models. When considering the magnitude of AUC above 0.5 (i.e. the feature is randomly distributed in cases and controls) as the power of a feature in a predictive task, however, using laboratory tests as HPO terms still preserved 73.9% of the power compared to using laboratory tests as the original continuous values. Taken together, the analysis confirmed that transforming laboratory tests from the continuous numeric values into HPO-coded phenotypes still preserved the majority of information.

**Figure 7.**
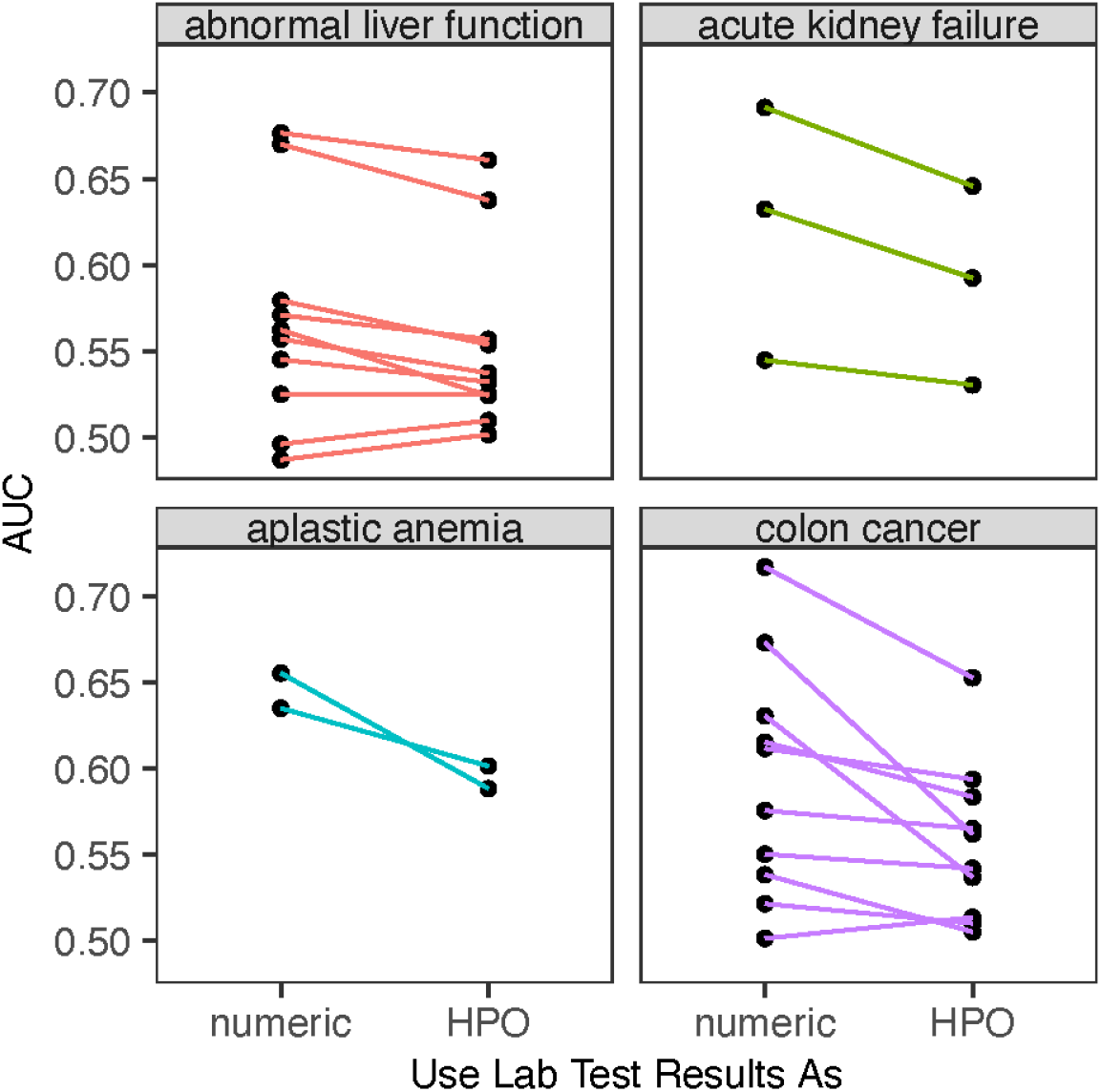
Performance of laboratory tests in predictive tasks when used as the numeric values vs HPO terms.

## Discussion

In this report, we presented building patient phenotypic journeys from clinical laboratory tests in a real-world EHR dataset after the LOINC2HPO transformation. Among 1.07 billion laboratory test records in the Sema4 Data Warehouse, we successfully converted 774 million (72.5%) into HPO-coded phenotypes. The high conversion rate confirms that LOINC2HPO can be successfully used to analyze laboratory data stored in the tabular format within relational databases even though it was initially developed for the Fast Healthcare Interoperability Resources (FHIR) format ^12^. The transformation allowed us to describe each patient with medically relevant phenotypes and thereby create patient phenotypic journeys throughout their healthcare history. Global analysis of the patient phenotypic journeys revealed longitudinal changes of both tested phenotypes and observed phenotypes. Furthermore, we found clear racial patterns especially in what phenotypes were tested from laboratory test orders. The vast collection of phenotypic journeys also allowed us to screen for what abnormal phenotypes correlated with future asthma progression into the severe form. Lastly, the transformation of laboratory test results into HPO-coded phenotypes caused a reduction of performance in predictive tasks but nevertheless preserved the majority of information.

A concern for transforming the continuous values of laboratory tests into HPO-coded phenotype terms is loss of information. Many previous work using laboratory tests for translational research had relied on feeding statistical models with the continuous values ^31,32^. A previous work also discovered that the variations of laboratory test values within the reference range, even if the test result falls within the range, have medical significance ^33^. In this report, we quantified the loss of performance in predictive tasks and our own result confirmed the expected loss of information by 26.1% (Figure 7). One unique advantage of LOINC2HPO, however, is to allow one to systematically transform laboratory tests into HPO-coded phenotypes that can describe patients with medically relevant terms. The approach achieves data integration on two levels. On the LOINC level, different LOINC codes are mapped to the same HPO terms if they all measure similar medical conditions, and therefore the laboratory test results will be mapped to the same set of HPO terms. On the HPO level, the hierarchical structure of HPO allows one to integrate granular phenotypes with the more generic ancestor terms. Therefore, the loss of information by 26.1% is an overestimation as we only utilized one laboratory test at a time for simplicity. Additionally, integrating different laboratory tests together can effectively increase sample size for specific medical phenotypes and allow statistical analysis that is otherwise difficult or impossible. We concluded that with limited information loss, the LOINC2HPO transformation generated deep phenotypes and opened the possibilities for many downstream applications.

Transforming laboratory tests into HPO-coded phenotypes allowed us to build a vast collection of patient phenotypic journeys for 2.2 million patients. Patient journey is a description of medical events that happen to a subject throughout the care history for a given condition ^1^. It has long been an interest for the pharmaceutical industry as they provide the foundation to uncover unmet medical needs, and thereby, opportunities for novel interventions. In this study, we created patient phenotypic journeys by summarizing how many abnormal phenotypes were observed from laboratory tests each year and how many were ruled out by laboratory tests that yielded within-range values. For practical uses, patient journeys can be customized so that the time window is in relative to the medical outcome under study and with a larger or smaller unit length (e.g. 3-Year, or 1-Month). Additionally, there were previous efforts in mapping diagnosis codes into HPO terms ^8^, or text mining HPO from imaging studies or doctor notes ^7,34^. We envision future patient journeys to include those EHR data sources for building comprehensive patient phenotype journeys as laboratory tests are heavily biased toward human physiology that is easily measured by highly automated equipment.

A global analysis of patient journeys revealed longitudinal changes and racial differences in what phenotypes were observed or tested in the dataset. We found an increased proportion of patients being confirmed to have Abnormality in the genitourinary system [HP:0000119], which is consistent with recent findings of globally increased urinary tract infection ^35^ and end stage renal disease ^36^ in the past three decades. However, we also caution that the current dataset is mainly from a regional hospital system and that the trends may not always be representative on a national or global level. Because laboratory tests can be used for both differential diagnosis and preventative screening, there are also two competing explanations for any longitudinal trends or racial patterns, either they reflect a medical necessity for diagnosis purposes or how frequently they were used for preventative screenings. For example, our discovery that African American were more likely to be tested for CD4+ T helper cells, is probably due to the established medical consensus that this population has higher HIV burden ^19^. On the contrary, the increased testing for abnormality of the nervous system and the endocrine system (Figure 5) is possibly for screening purposes as there is an increased awareness in the general public for mental health issues.

Patient phenotype journeys can also be combined with other EHR data sources for translational research. In this study, we used severe asthma progression as an endpoint. We utilized logistic regression, a commonly used statistical model in medicine, to establish whether each abnormal phenotype in a given time window was correlated with the endpoint. One notable difference from our previous study of LOINC2HPO ^12^, is that we created two features for each abnormal phenotype, whether the phenotype was tested and whether the phenotype was observed. The fact that we were able to identify Eosinophilia and Neutrophilia as two of the most significant phenotypes associated with severe asthma progression validated the approach and the usefulness of our patient journeys. Sputum eosinophil and neutrophil counts were established as golden criteria to diagnose and subtype severe asthma ^26^. Our findings provided evidence that routine laboratory tests with peripheral blood that were performed up to three years prior were also statistically significantly correlated with severe asthma progression. We anticipate future patient phenotypic journeys that include data from other EHR sources, including imaging and clinical notes, to serve as an even more robust resource for biomarker screens in many therapeutic areas.

## Data Availability

Due to patient privacy protection, the dataset is not publicly accessible. Data access requests
should be addressed to Sema4 (https://support.sema4.com/hc/en-us/requests/new).

## Supplemental Material

Supplemental Figures can be accessed at

https://docs.google.com/document/d/1aW2mJNzBNLUD3jG8y3A2tcfjjr82qv3goAOhTZc3PjI/edit?usp=sharing.

## Code Availability

Code for data engineering are available at https://github.com/sema4hai/s4-msdw2trinetx, for data analysis: https://github.com/sema4hai/s4-loinc2hpo-msdw, and for the interactive web viewer: https://github.com/sema4hai/s4-loinc2hpo-phenoviewer.

## Data Availability

Due to patient privacy protection, the dataset is not publicly accessible. Data access requests should be addressed to Sema4 (https://support.sema4.com/hc/en-us/requests/new).

## Acknowledgement

The authors acknowledge Yun Mai (Sema4) and Rene Dempsey (Sema4) for their effort in mapping laboratory test code from the local codeset to LOINC; Kalpana Raja (The University of Texas Health Science Center) for editing the manuscript; Hua Xu (The University of Texas Health Science Center) for reviewing the manuscript and providing constructive suggestions. P.N.R. is funded by National Institute of Health (NIH). The content is solely the responsibility of the authors and does not necessarily represent the official views of the National Institutes of Health, nor should any endorsements be inferred by NIH or the U.S. Government.

## Contributions

X.A.Z.: data engineering, data analysis and interpretation; K.L., L.J., Z.L., L.A., J.T.: data analysis and interpretation; H.X., W.O., M.H., Q.P., G.S., E.S., P.N.R.: interpretation; X.W.: data analysis, interpretation. X.A.Z., X.W.: designed study, wrote manuscript; X.W.: supervised study. All authors contributed to manuscript revisions.

## Conflict of interest statements

X.A.Z., K.L., L.J., Z.L., L.A., T.J., M.K.H., Q.P., W.O., G.S., E.S., and X.W. are employed by Sema4 OpCo, Inc..

## Ethics committee approval

The study is exempt from review by the Institutional Review Board (IRB) because the data is de-identified.

## Notes

### Funding Statement

The study was funded by Sema4 OpCo, Inc. P.N.R. is funded by National Institute of Health (NIH). The content is solely the responsibility of
the authors and does not necessarily represent the official views of the National Institutes of
Health, nor should any endorsements be inferred by NIH or the U.S. Government.

### Author Declarations

Ethics committee/IRB of Sema4 OpCo, Inc waived ethical approval for this work.

