## Supplemental Materials for "Building Population Phenotypic Journeys from Laboratory Tests in Electronic Health Records for Translational Research"

A

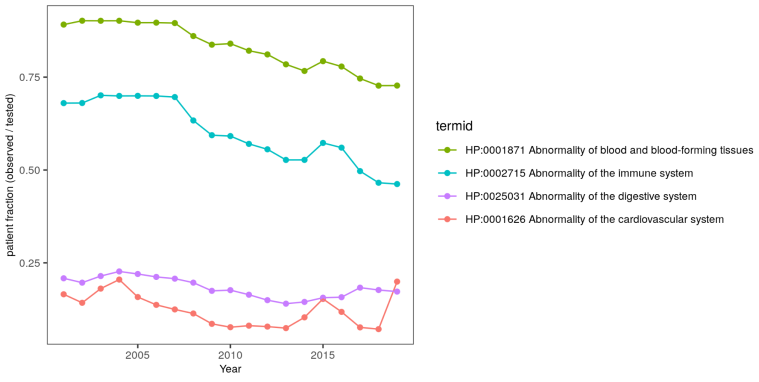


B

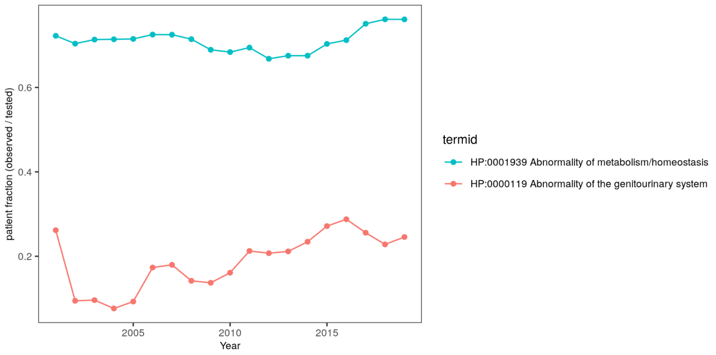


Supplemental Figure 1. Patient count with an observed phenotype normalized to how many patients were actually tested. A. Plot for 4 phenotypes of which the patient count decreased when normalized to database size (see Figure 4B). B. Plot for 2 phenotypes of which the patient count increased when normalized to database size (see Figure 4B).

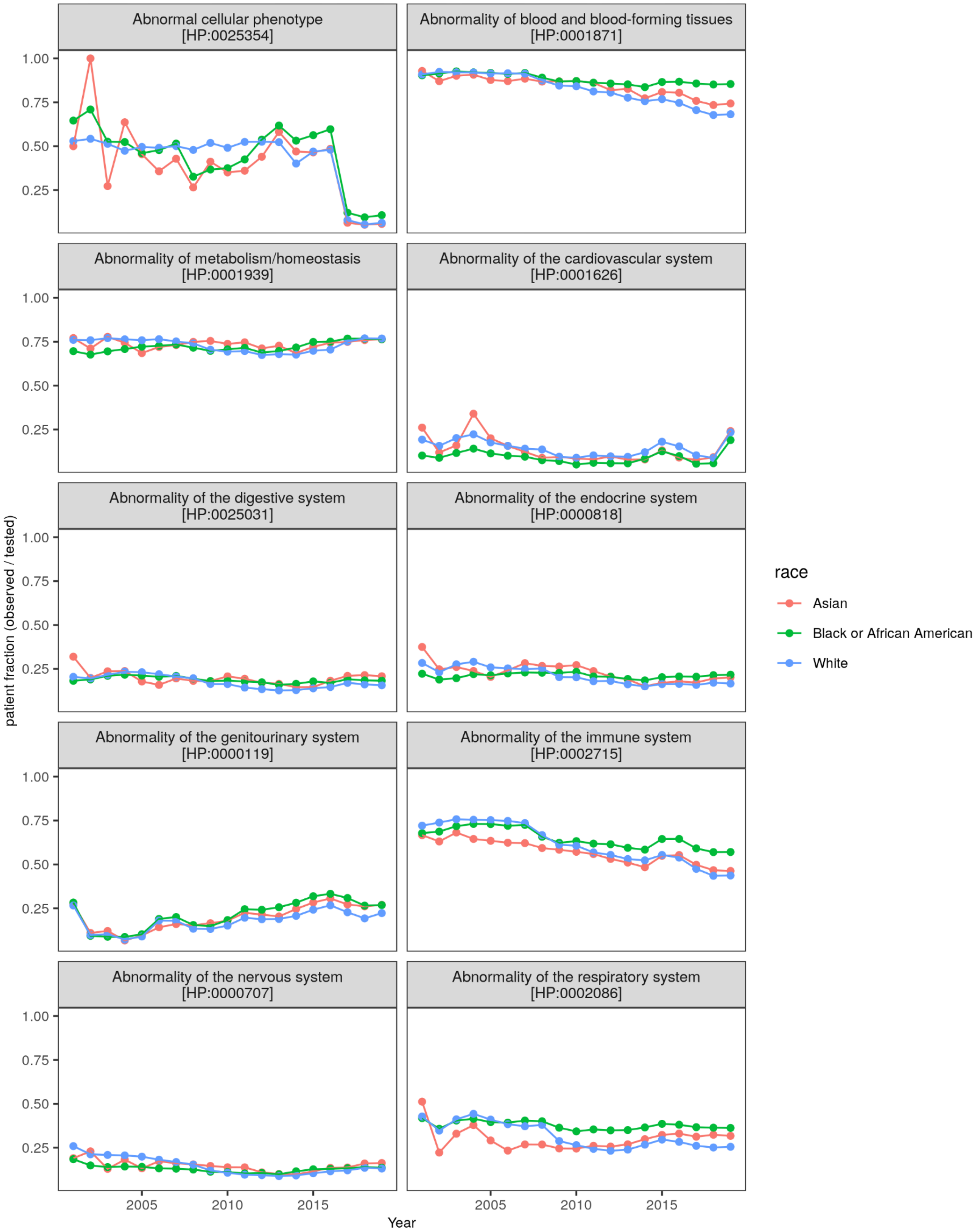


Supplemental Figure 2. Fraction of patients observed for a phenotype among those that were tested. In each calendar year, we counted the number of patients who were observed to have a specific phenotype and then normalized it by the total number of patients tested for the phenotype. Only high level HPO terms were shown in the figure.

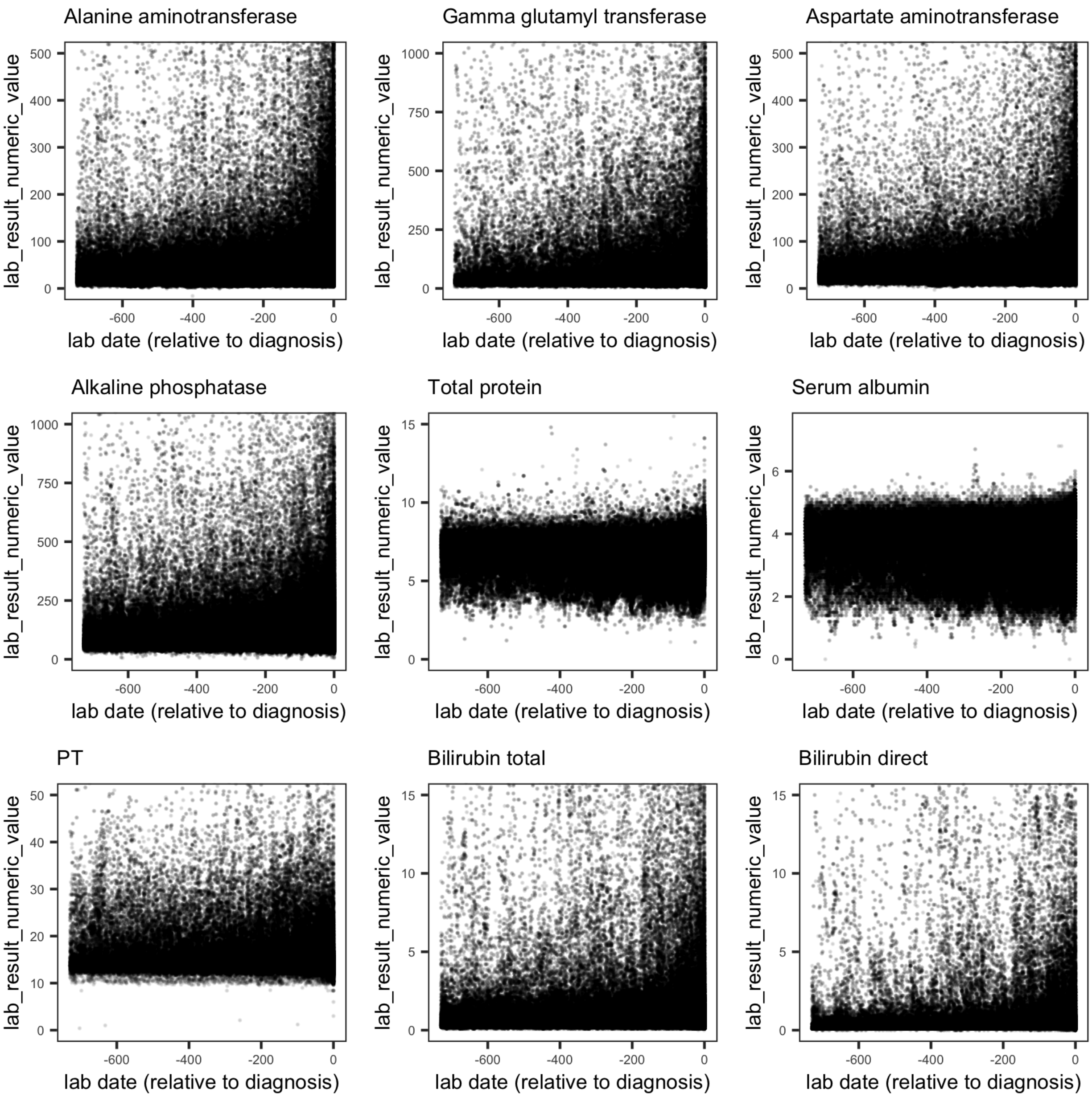


Supplemental Figure 3. Reported laboratory test values prior to the diagnosis of abnormal liver function. All the patients have at least one diagnosis of abnormal liver function. We determined the earliest date of abnormal liver function diagnosis, and then we collected all the laboratory test values. We plotted the laboratory test values (y-axis) against the relative time to the first diagnosis (x-axis, in days). Each subpanel represents one laboratory test as indicated above the plot. PT: prothrombin time.

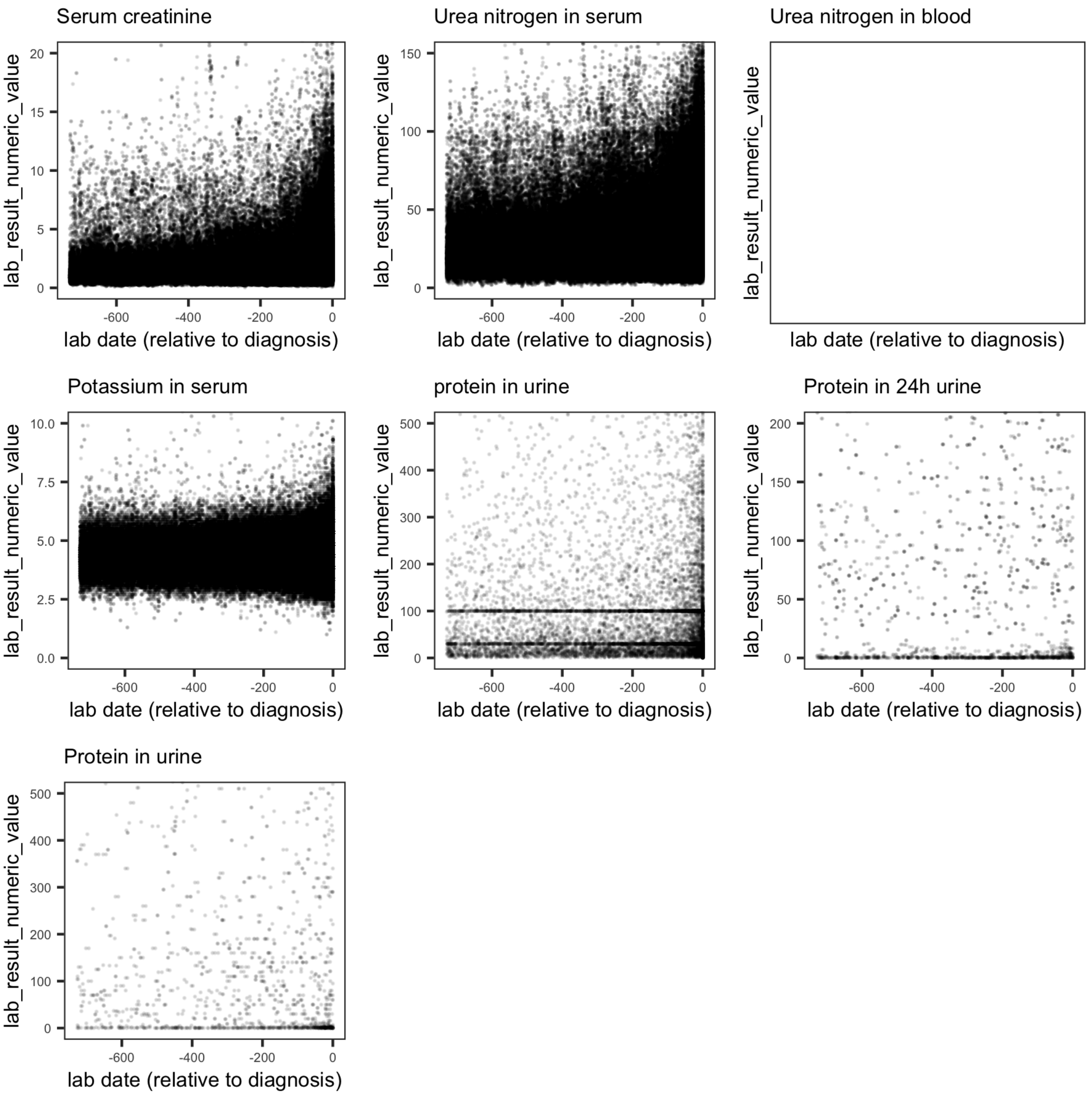


Supplemental Figure 4. Reported laboratory test values prior to the diagnosis of acute kidney failure. All the patients have at least one diagnosis of acute kidney failure. We determined the earliest date of acute kidney failure diagnosis, and then we collected all the laboratory test values. We plotted the laboratory test values (y-axis) against the relative time to the first diagnosis (x-axis, in days). Each subpanel represents one laboratory test as indicated above the plot. Empty panel indicates data are missing.

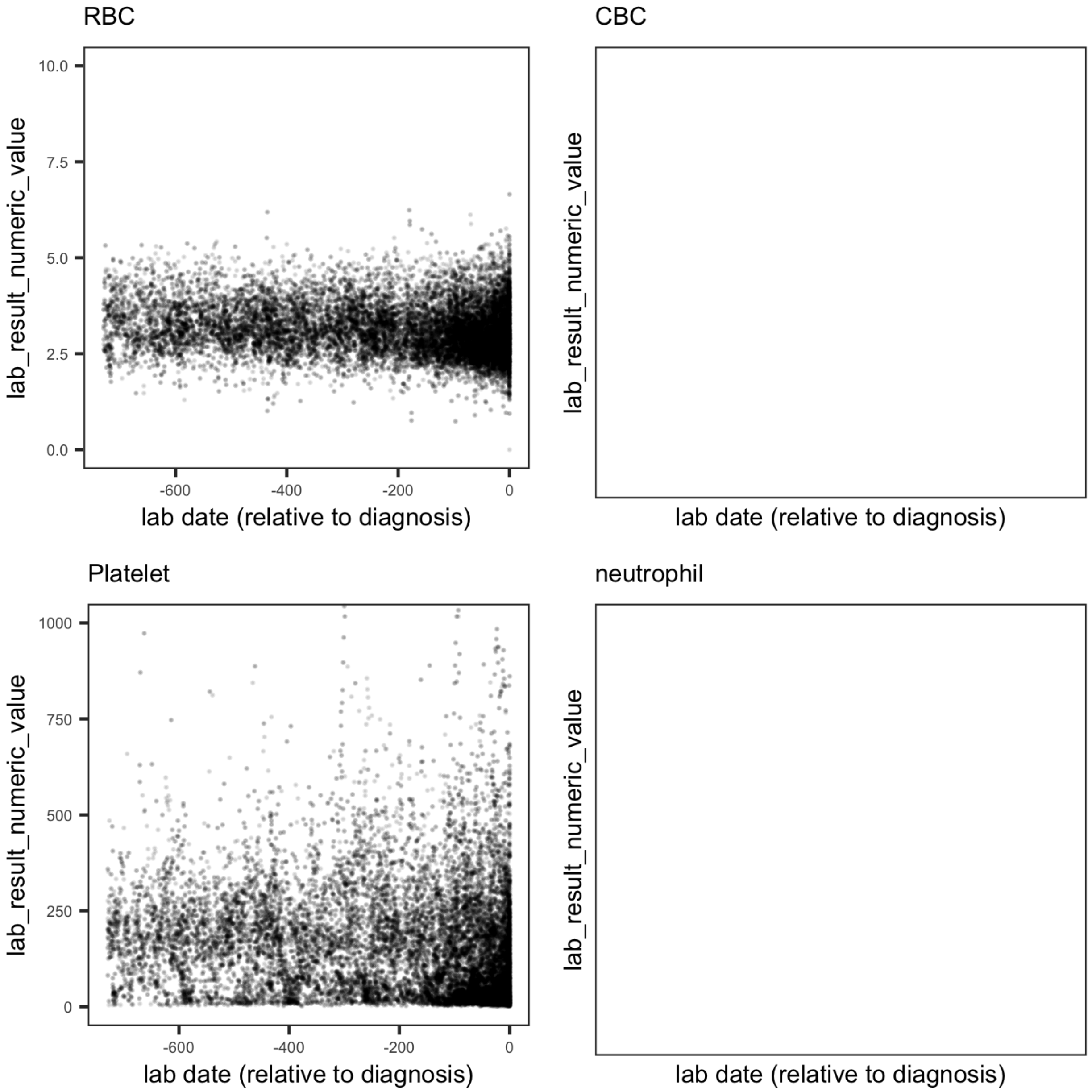


Supplemental Figure 5. Reported laboratory test values prior to the diagnosis of aplastic anemia. All the patients have at least one diagnosis of aplastic anemia. We determined the earliest date of aplastic anemia diagnosis, and then we collected all the laboratory test values. We plotted the laboratory test values (y-axis) against the relative time to the first diagnosis (x-axis, in days). Each subpanel represents one laboratory test as indicated above the plot. RBC, red blood cell count; CBC, complete blood cell count. Empty panel indicates data are missing.

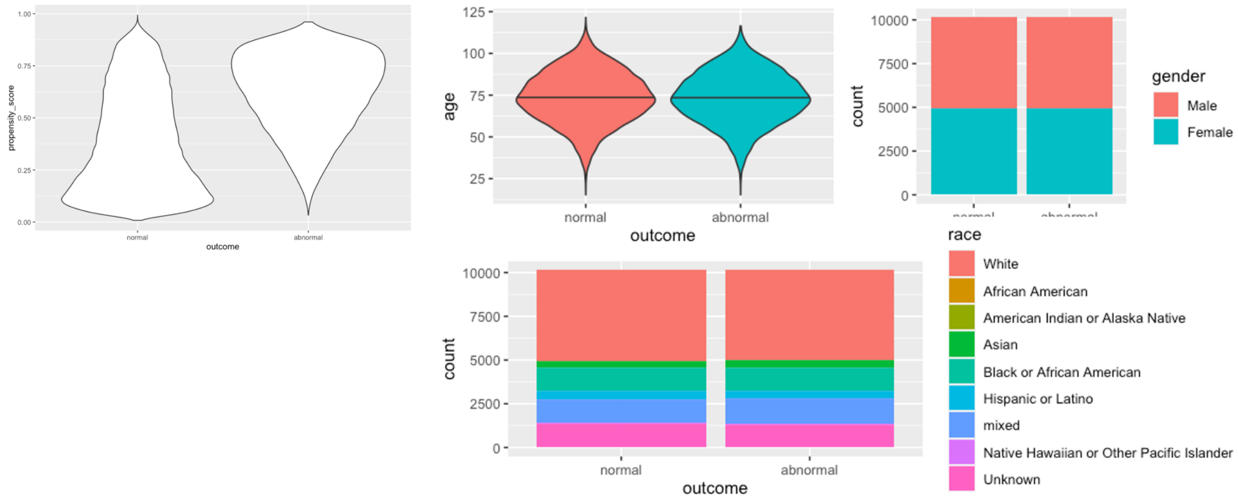


Supplemental Figure 6. Patient matching for colorectal cancer cases and controls based on propensity score. A. Distribution of calculated propensity score for colorectal cancer based on age, sex and race. Normal, cancer free (i.e. control); abnormal, colorectal cancer (i.e. case). B, C, D shows the distribution of age, sex and race after subsampling cancer free patients to find matched controls.

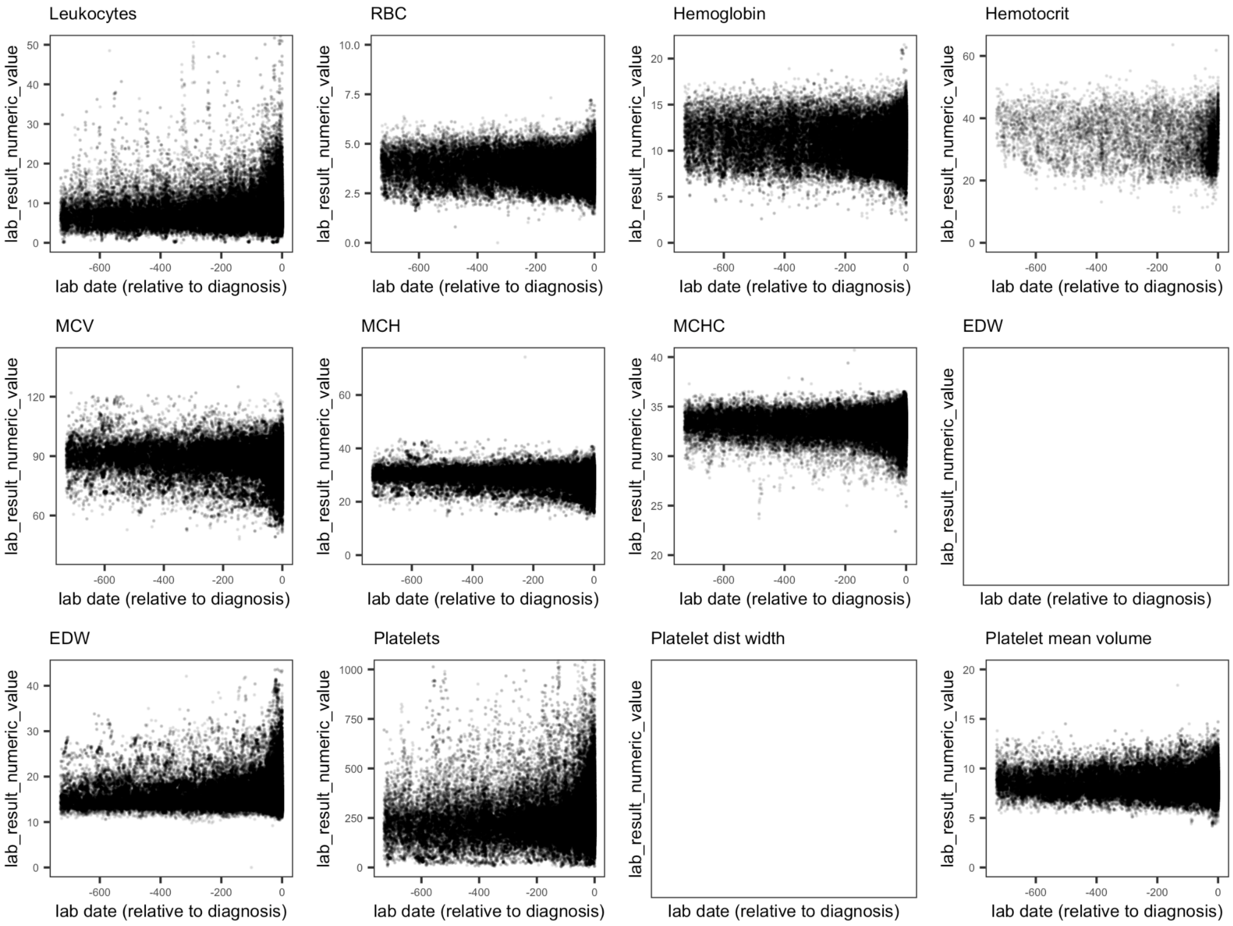


Supplement Figure 7. Reported test values for colorectal cancer and matched controls. All the patients have at least one diagnosis of colorectal cancer. We determined the earliest date of aplastic colorectal cancer, and then we collected all the laboratory test values. We plotted the laboratory test values (y-axis) against the relative time to the first diagnosis (x-axis, in days). Each subpanel represents one laboratory test as indicated above the plot. RBC, red blood cell count; MCV, mean corpuscular volume; MCH: mean corpuscular hemoglobin measurement; MCHC: mean corpuscular hemoglobin concentration; EDW: erythrocyte distribution width. Empty panel indicates data are missing. Two LOINC codes were collected for EDW, 21000-5 (Erythrocyte distribution width [Entitic volume] by automated count, data missing) and 788-0 (Erythrocyte distribution width [Ratio] by Automated count, data not missing).
